# Off-label evaluation of the BD MAX MDR-TB assay for rapid diagnosis of rifampicin and isoniazid resistance of *Mycobacterium tuberculosis* clinical isolates in a high-volume reference laboratory

**DOI:** 10.1101/2025.06.17.25329767

**Authors:** Angela Pires Brandao, Fabiane Maria de Almeida Ferreira, Fernanda Cristina dos Santos Simeao, Lucilaine Ferrazoli, Erica Chimara, Rosângela Siqueira de Oliveira, Juliana Maira Watanabe Pinhata

## Abstract

Drug-resistant tuberculosis (TB) remains a major global health concern. Multidrug-resistant TB is defined by resistance to at least rifampicin (RIF) and isoniazid (INH), the two key drugs used in TB treatment. The BD MAX™ Multi-Drug Resistant Tuberculosis (BD MAX) assay is a fully automated real-time PCR platform, recommended by the World Health Organization for the initial diagnosis of TB and RIF and INH resistance (RIF-R and INH-R) directly from pulmonary clinical samples. This study aimed to assess the off-label performance of BD MAX in clinical *M. tuberculosis* complex (MTBC) isolates under routine laboratory conditions. The assay was first validated using non-tuberculous mycobacteria (NTM) and MTBC isolates with known mutations. For real-world validation, it was compared to the GenoType MTBDR*plus* by testing 1,440 clinical isolates prospectively. The BD MAX assay correctly excluded MTBC from all NTM cultures. Among MTBC isolates with known mutations, it identified 19 of 20 RIF-R isolates and 14 of 15 INH-R isolates. In prospective testing, BD MAX achieved 99.8% sensitivity (1,406/1,409), 100% specificity (31/31), and 99.8% (1,437/1,440) overall accuracy for MTBC detection. For drug resistance detection, it showed 95.2% (40/42) concordance for RIF, 96.8% (30/31) for INH and 81.3% (13/16) for MDR when compared to MTBDR*plus*. Discrepancies between MTBDR*plus* and BD MAX included heteroresistant cases and unreportable resistance results by BD MAX due to infrequent mutations or low bacterial load. Overall, this study confirms BD MAX as an accurate and reliable tool for MTBC detection and drug resistance profiling in clinical isolates in high-volume TB laboratories.

**Importance:** This study highlights the importance of the BD MAX™ Multi-Drug Resistant Tuberculosis assay (BD MAX) applied in clinical isolates for the detection of multidrug-resistant tuberculosis (MDR-TB), i.e., *Mycobacterium tuberculosis* resistance to rifampicin and isoniazid. TB is a global health issue, and drug-resistant TB makes treatment more difficult, favoring transmission and disease amplification. The BD MAX platform offers a faster and more automated way to detect TB and drug resistance. The study showed that BD MAX applied off-label in clinical isolates accurately identified TB and resistance to rifampicin and isoniazid, with results comparable to the widely used line probe assay. This is significant in a high-volume laboratory because it is simpler and more rapid than the line probe assay. BD MAX showed some limitations, especially in detecting rare mutations and in cases of low bacteria levels. Overall, this tool could improve TB care, especially in high-volume laboratories.

## Introduction

According to the World Health Organization (WHO), in 2023, only two out of five of the estimated 400,000 new cases of rifampicin-resistant (RIF-R-) or multidrug-resistant tuberculosis (MDR-TB) were diagnosed and received appropriate drug-resistant TB treatment(1). This is concerning, as effective TB management and control rely on the prompt diagnosis of the disease and *Mycobacterium tuberculosis* complex (MTBC) drug resistance, along with the timely initiation of an effective treatment regimen(2). To address this issue, the WHO recommends universal drug susceptibility testing (DST), advising that a rapid initial test for resistance to RIF should be conducted for all individuals suspected of having TB(3).

Brazil is one of the 30 countries with high burdens of TB and TB/HIV coinfection, making it a priority for TB control efforts(3). In 2023, Brazil reported over 80,000 new TB cases, and there were 5,800 deaths attributed to the disease in 2022(4). The state of São Paulo contributes significantly to this burden, accounting for approximately 25% of the new TB cases reported annually in the country. In 2023, São Paulo recorded 19,571 new TB cases, resulting in an incidence rate of 42 per 100,000 residents. The state also reported 1,283 deaths from TB in the same year, with a mortality rate of 2.8 per 100,000 inhabitants. In terms of drug resistance, there were 182 new cases of drug-resistant TB in São Paulo in 2023, including 118 cases of RIF-R-or MDR-TB(4), which is defined as MTBC resistance to at least RIF and isoniazid (INH).

Molecular tests for drug-resistant TB, which detect mutations affecting genes or the expression of genes associated with drug resistance, are both rapid and highly accurate. These tests offer high sensitivity and specificity(5), enabling rapid diagnosis and the administration of effective treatment to patients(6).

Currently, in Brazil, the most commonly used commercial molecular test for detecting drug-resistant TB directly from clinical specimens is the Xpert® MTB/RIF Ultra (Cepheid, USA)(7). While this test is fully automated, its throughput is limited because the available equipment in Brazil can analyze only four samples at a time. Additionally, this test does not detect INH monoresistance, which is present in 8% of TB cases globally(8). The Xpert® MTB-XDR (Cepheid), which identifies resistance to INH and second-line drugs, is not yet in use in the country.

In Brazil, the line probe assays (LPAs) GenoType® MTBDR*plus* and GenoType® MTBDR*sl* version 2.0 kits (Hain Lifescience-Bruker, USA) are used alongside the automated BACTEC MGIT 960 phenotypic DST (pDST) (Becton, Dickinson and Company, USA) for detecting MTBC drug resistance to first– and second-line TB drugs in some states’ reference laboratories(9). However, pDST is time-consuming and carries significant biosafety risks. Additionally, performing LPAs requires specialized training and a complex infrastructure, with distinct areas designated for the various procedures involved in the tests(7).

In recent years, several automated molecular assays have been developed for the detection of MTBC and its resistance to RIF and INH using centralized platforms, which can also diagnose other infectious diseases, and thus are suitable for implementation in reference laboratories. One such assay is the BD MAX™ Multi Drug Resistant Tuberculosis (BD MAX MDR-TB) (Becton, Dickinson and Company), herein referred to as BD MAX. This test identifies MTBC-specific DNA sequences (insertion sequences *IS6110* and *IS1081*, in addition to a single copy target). It also detects mutations associated with resistance to RIF (in the RIF resistance determining region of the *rpoB* gene, RRDR, codons 507-533) and INH (in the *inhA* gene promoter region and *katG* gene codon 315)(10).

The BD MAX is an integrated assay that automates the entire process of MTBC lysis, DNA extraction, and detection of molecular targets through real-time PCR. It is designed for direct detection from untreated sputum samples or sediment from decontaminated sputum samples. The assay can process up to 24 samples per run and delivers results within four hours(11,12).

Centralized platforms such as BD MAX allow the diagnosis to be performed with high accuracy for a large number of samples processed simultaneously. Its automated design minimizes testing errors and reduces biosafety risks associated with handling respiratory specimens(7). This makes the BD MAX well-suited for reference laboratories with high testing volumes, such as the Núcleo de Tuberculose e Micobacterioses at the Adolfo Lutz Institute in São Paulo. This reference public health laboratory handles the identification and drug resistance detection of over 6,000 MTBC isolates annually, which are received from the state’s TB network laboratories, using the LPAs GenoType MTBDR*plus* and MTBDR*sl* v. 2.0.

Integrating the BD MAX for clinical isolates into the routine for diagnosing MDR-TB would enable a faster and more automated process compared to the LPAs, which require numerous manual steps and are prone to cross-contamination. Additionally, BD MAX requires fewer separate rooms in the laboratory, simplifying its implementation. Therefore, the aim of this study was to assess the off-label performance of the BD MAX assay when applied to clinical MTBC-suggestive isolates in the routine operations of a reference laboratory, compared to the LPA GenoType MTBDR*plus*.

## Materials and Methods

### Retrospective validation of the BD MAX MDR-TB assay

First, the BD MAX assay was validated on culture samples regarding its limit of detection (LoD), the optimal volume of inactivated culture samples to be processed and the time of incubation of these samples with the STR reagent, using the reference strain *M. tuberculosis* H37Rv (ATCC 27294). After these parameters were established, non-tuberculous mycobacteria (NTM) isolates and MTBC clinical isolates with known mutations were tested on the BD MAX. This initial validation was performed before the assay was conducted prospectively on the cultures routinely subjected to the LPA.

### Culturing, inactivation and DNA extraction of the isolates

Isolates and *M. tuberculosis* H37Rv stored at –70°C in the laboratory repository were reactivated in Löwenstein-Jensen (LJ) or MGIT at a biosafety level 3 (BSL-3) laboratory, for BD MAX validation. The cultures were inactivated for testing in the BD MAX system, as the STR reagent was designed for the pre-treatment of sputum samples, not being effective in inactivating the high number of bacilli typically found in cultures. The inactivation was also necessary because the BD MAX equipment was located in an area outside the BSL-3 laboratory. One mL of the homogenized growth of the cultures in MGIT or a loopful of the growth on LJ diluted in 1 mL of ultrapure water was incubated at 95°C for 20 minutes for inactivation. To enhance cell lysis for DNA extraction by BD MAX, these heat-inactivated cultures were frozen, incubated again at 95°C in a thermoblock for 20 minutes(13), and stored at –20°C.

### Assessment of the LoD of the BD MAX MDR-TB assay

BD MAX assay’s LoD on culture samples was assessed using *M. tuberculosis* H37Rv grown for 28 days at 35±2°C in LJ. To prepare bacterial suspensions for testing, first, a 10 µL loopful of the strain growth was emulsified along the inside wall of a sterile tube. The emulsified colonies were then placed in a sterile screw-cap glass tube containing 5-10 sterile 3 mm glass beads. After securely closing the cap, the tube was vortexed for at least 2 minutes until the clumps were well dispersed. Subsequently, 5 mL of fresh sterile distilled water was added and the tube’s content was homogenized by vigorous vortexing for an additional 2 minutes. The tube was then left undisturbed for 30 minutes to allow remaining clumps to settle. The turbidity of the supernatant was adjusted to McFarland 0.5 in a new sterile glass tube using sterile distilled water and the suspension was vortexed for 30 seconds. The turbidity of the suspension was measured using a densitometer(14).

A 1:100 dilution of the bacterial suspension was then prepared in 7H9/OADC broth through two tenfold dilution steps. First, a 10⁻¹ suspension was prepared by adding 1 mL of the 0.5 McFarland bacterial suspension to 9 mL of 7H9/OADC broth and vortexing until well mixed. From this, a 10⁻² suspension was prepared by adding 1 mL of the 10⁻¹ suspension to 9 mL of 7H9/OADC broth and vortexing again. Additionally, from the 10⁻² suspension, a 10⁻^3^ suspension was prepared, and so on until obtaining a 10^−8^ suspension. The 10⁻¹ suspension contained 1 × 10^6^ colony forming units (CFU)/mL, the 10^−2^ contained 1 × 10^5^ CFU/mL(14), and so on, until the 10^−8^ suspension, which contained no CFU. All the suspensions were inactivated as described above before undergoing BD MAX testing.

Since we used an H37Rv culture rather than raw sputum samples, which are more difficult to fluidize, and were evaluating the use of the BD MAX in mycobacterial cultures, we also investigated whether the 25-min incubation after the first room temperature 5-min incubation of the inactivated H37Rv culture + STR mixture (as recommended by the manufacturer for sputum samples) would impact the performance of the assay. Additionally, considering the high number of isolates received by our laboratory, we also tested whether incubating the mixture of H37Rv culture + STR for 24, 48, and 72h would affect the BD MAX performance (Table 1). For this, 1 mL of each inactivated suspension of H37Rv (10^−1^ to 10^−8^) was added to 2 mL of STR, and 2.5 mL of this mixture was tested in the BD MAX.

**Table 1.**
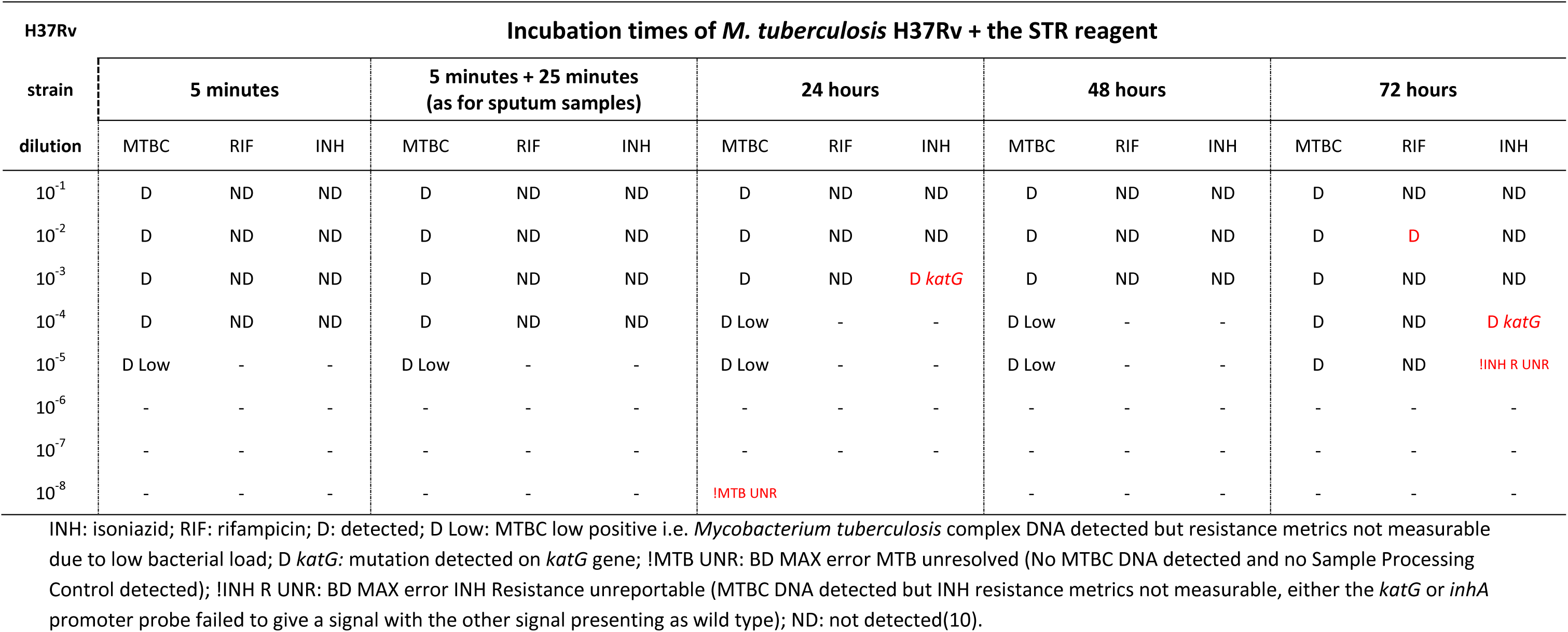
Results of dilutions and incubation periods of the H37Rv reference strain with STR reagent for the assessment of the limit of detection of the BD MAX MDR-TB assay for off-label detection of *Mycobacterium tuberculosis* clinical isolates.

The LoD of the BD MAX assay for MTBC detection was 10^−5^, corresponding to 100 CFU/mL, for all H37Rv + STR incubation protocols (5-min, 5-min + 25-min, 24h, 48h, and 72h) (Table 1). For drug resistance mutations detection, the LoD was 10^−4^ (corresponding to 1,000 CFU/mL) for the 5-min and 5-min + 25-min protocols. For the 24h, 48h and 72h incubation protocols, false-positive mutation results were obtained. Therefore, we used the 5-min incubation protocol for all BD MAX tests due to its faster processing time.

### Standardization of the inactivated culture samples for the BD MAX MDR-TB assay

We receive mycobacterial cultures on solid media or liquid MGIT media from 82 TB laboratories across the state. However, we do not have information about the age of the cultures upon their arrival at our laboratory. Additionally, some cultures arrive with very sparse growth, particularly those in MGIT, as the BACTEC equipment is highly sensitive in detecting even minimal growth. Similarly, older cultures on solid media often present with only a few colonies. Furthermore, even cultures with little growth present a significantly higher concentration of bacilli than clinical samples. Consequently, using the standard 2:1 ratio of STR to culture—recommended for clinical samples—may inhibit the BD MAX qPCR reaction. Thus, to standardize the volume of the cultures for testing on the BD MAX, we evaluated an inactivated H37Rv culture grown in MGIT for 28 days, using various ratios of culture to STR: 100 µL of culture to 2.9 mL of STR, 200 µL of culture to 2.8 mL of STR, 300 µL of culture to 2.7 mL of STR, and continuing to the maximum of 1 mL of culture to 2 mL of STR. As we obtained valid results for all ratios tested (data not shown), we decided to use the proportion of 200 µL of inactivated culture to 2.8 mL of STR (1:15) for all the subsequent tests. These volumes (resulting in a total of 3 mL) were used because 2.5 mL of the sample + STR mixture must be added to the BD MAX reaction tube.

We also evaluated whether adding STR would impact the performance of the BD MAX assay on the cultures, since the STR was not necessary to kill the already inactivated isolates. To do this, three MTBC clinical isolates with known mutations from our repository grown in MGIT were tested in duplicate. For each isolate, one tube contained the inactivated suspension mixed with ultrapure water, and the other tube contained the inactivated suspension mixed with STR. The proportion used was 200 µL of the suspension to 2.8 mL of water or STR. We observed a significant decrease in cycle threshold (Ct) values (by 9–10 cycles) when using STR for all three clinical isolates with known mutations. Therefore, we concluded the use of STR was necessary for a better performance of the BD MAX tests.

### BD MAX MDR-TB testing with non-tuberculous mycobacteria (NTM) and MTBC clinical isolates with known mutations

To assess the specificity of the BD MAX assay on culture samples, the following NTM strains from our repository were tested: *M. abscessus abscessus*, *M. abscessus bolletii*, *M. abscessus massiliense*, *M. avium*, *M. chelonae*, *M. fortuitum*, *M. intracellulare*, *M. kansasii*, and *M. peregrinum*. A clinical isolate identified as *Nocardia asteroides* was also tested.

To evaluate the BD MAX for drug resistance detection in clinical isolates, 29 MTBC isolates from our repository were used. These isolates have been previously characterized for phenotypic and molecular resistance to RIF and INH. Selection was based on mutations identified by either GenoType MTBDR*plus* or DNA sequencing (Table 2).

**Table 2.**
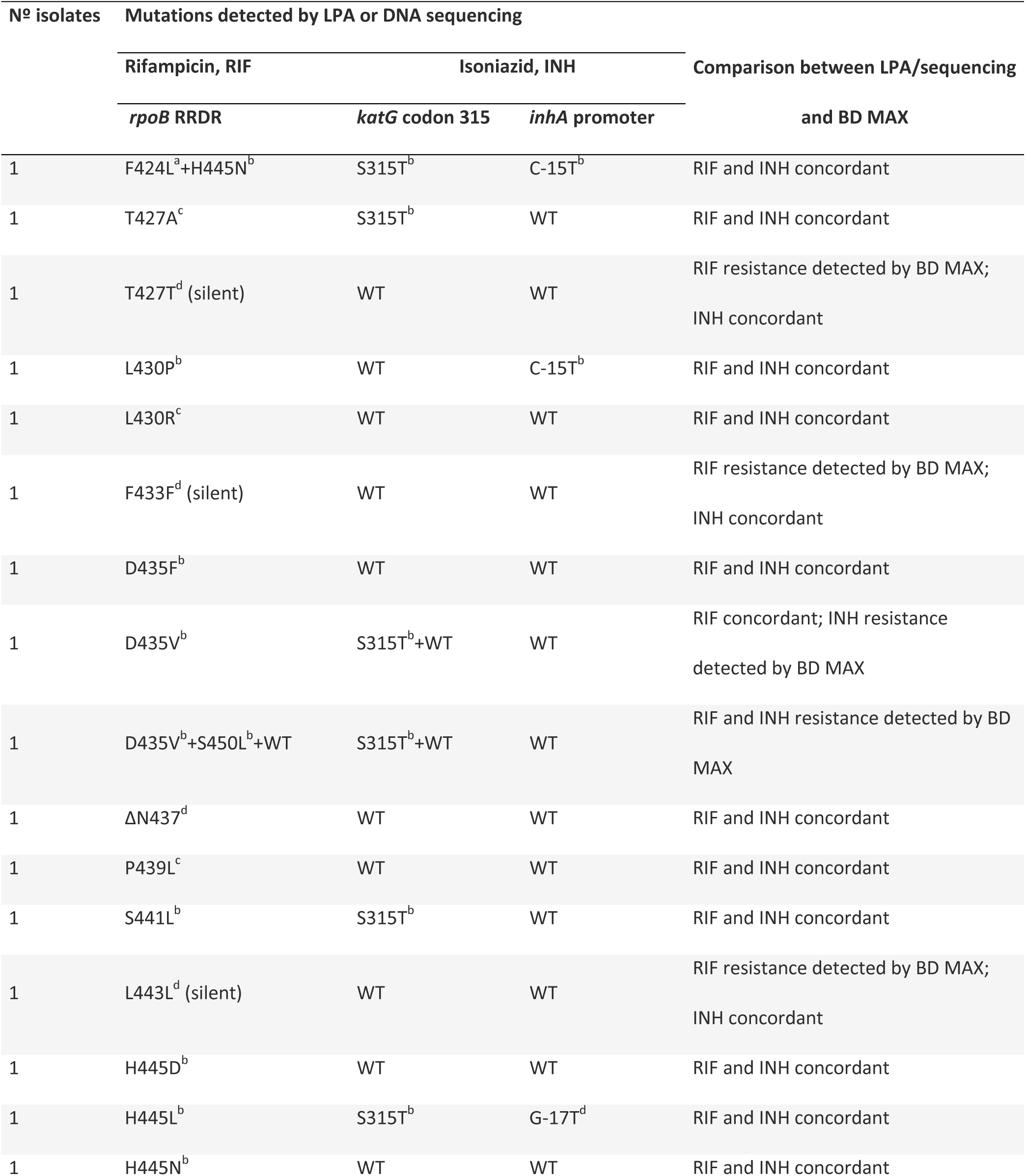
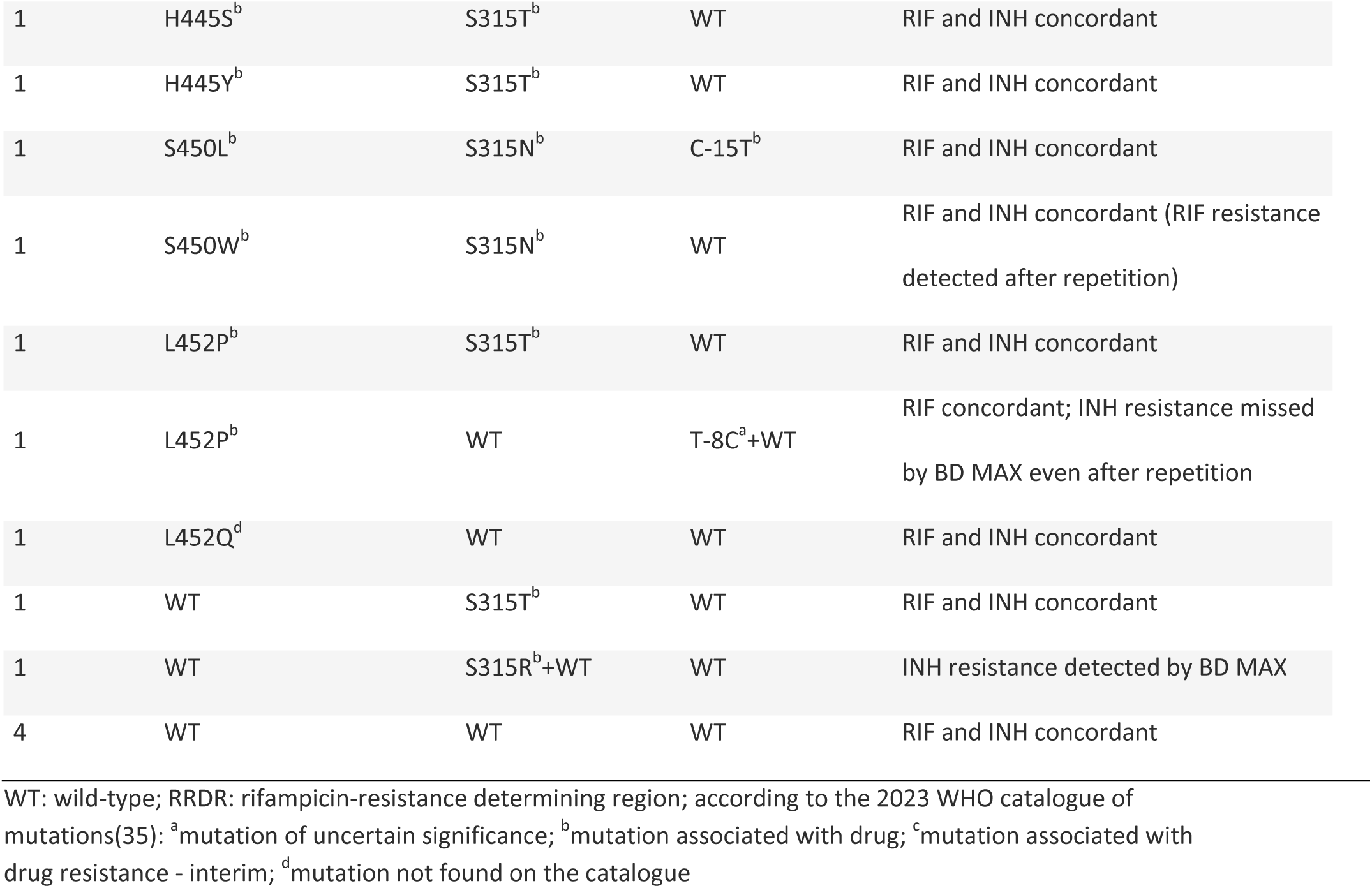
Clinical isolates of the *Mycobacterium tuberculosis* complex tested retrospectively by the BD MAX MDR-TB assay.

### Prospective testing on suggestive-MTBC clinical isolates by the GenoType MTBDR*plus* and BD MAX MDR-TB

#### Sample size calculation

Data from studies using the GenoType MTBDR*plus* and our laboratory’s testing production were used to calculate the sample size needed to evaluate the performance of BD MAX prospectively in our setting.

Applying the formula N ≥ Z^2^ P (1-P) / D^2^ [where N is the number of drug-resistant samples; Z = 1.96 for a 95% confidence interval; P is the expected sensitivity of the test (0.89 for INH; 0.93 for RIF), and D is the precision or confidence interval (0.1)](15), 38 INH-resistant and 25 RIF-resistant isolates were required.

Considering: (i) the resistance frequencies to RIF (1.8%) and INH (3.5%) in MTBC isolates tested by MTBDR*plus* in our setting in 2019; (ii) the 3.1% rate of MTBC-negative or invalid tests; and (iii) a 5% loss ratio; we estimated that 1,423 MTBC-suggestive cultures had to be tested to yield 25 RIF-R isolates. Given the higher frequency of INH resistance, this number of samples would also be sufficient to obtain 38 INH-R isolates.

#### Clinical isolates

The mycobacterial isolates are sent to our laboratory for species identification and DST by the TB network laboratories that perform culturing in the state of São Paulo. These isolates are first assessed regarding colony characteristics to presumptively determine whether they belong to the MTBC or are NTMs. When cultured on solid media, MTBC typically forms cream-colored, rough, and dry colonies. In liquid cultures, MTBC pure growth appears as cream-colored flakes, while the culture medium remains clear(16).

All clinical isolates presumptively identified as MTBC and tested with the GenoType MTBDR*plus* in our routine were prospectively included in the study. These isolates belonged to patients with criteria for DST as outlined by the State and National TB Control Programs(17). These criteria are: being a contact of a drug-resistant TB case; having a history of previous TB treatment; being HIV-positive or having other forms of immunosuppression; having a positive smear result at the end of the second month of treatment; belonging to high-risk populations for TB (e.g., healthcare workers, homeless individuals, prisoners, hospitalized patients, and indigenous people); or having a GeneXpert MTB/RIF Ultra RIF-R result.

#### DNA extraction

For DNA extraction, 1 mL of the isolate’s growth in MGIT was distributed into two vials each: one for MTBDR*plus* and the other for BD MAX. For isolates grown on solid media, a loopful of bacterial growth was added to two vials each, containing 1 mL of ultrapure water, with one vial designated for MTBDR*plus* and the other for BD MAX. DNA extraction for MTBDR*plus* was carried out according to the manufacturer’s instructions(18). The BD MAX vials were heat-inactivated and freeze-thawed as described above, and kept at –20°C until testing.

### GenoType MTBDR*plus*

This test was performed according to the manufacturer’s instructions(18). The hybridization of the PCR product with the nitrocellulose strips was performed using the GT-Blot 48 automated equipment (Bruker-Hain Lifescience), which allows the simultaneous processing of 48 tests(18). Once the strips had dried, the tray used for hybridization was placed into the GenoScan equipment (Bruker-Hain Lifescience). This equipment digitized the strips and transmitted the images to the GenoScan software (Bruker-Hain Lifescience). The software then interpreted the results based on the patterns of hybridized bands(18). After verification by the technician, the results were released into the laboratory system, following the routine process.

An inconclusive result was defined when the TUB band, indicating the presence of MTBC, was positive, but the WT bands for the genes were faint and there was no development of mutation (MUT) bands due to low bacterial load, thus impairing the detection of drug resistance. All isolates that yielded inconclusive results were retested.

Isolates that tested negative for MTBC by GenoType MTBDR*plus* were subsequently screened with the Bioline TB Ag MPT64 immunochromatographic test (Abbott, USA) for MTBC identification(19). Once the result was negative for MTBC, the isolates were tested with the GenoType Mycobacterium CM version 2.0 kit (Bruker-Hain Lifescience). NTM isolates that could not be identified to the species level by the GenoType Mycobacterium CM assay were further analyzed using the PRA-*hsp65* in-house methodology(20).

#### BD MAX MDR-TB assay

For BD MAX testing, 200 µL of inactivated isolates suspensions were added to 2.8 mL of the STR reagent. The mixture was shaken vigorously 10 times and incubated for 5 minutes at room temperature. Then, 2.5 mL of the mixture was transferred to the sample tube of the BD MAX kit and inserted into the equipment, following the manufacturer’s protocol(10).

The BD MAX equipment system automatically interpreted the results, which can be reported as: MTB detected/not detected/low positive (in the latter case, resistance is not analyzed), and RIF/INH resistance detected/not detected. A RIF/INH unreportable result means that MTBC DNA was detected but the resistance metrics for the drug could not be measured. An MTB unresolved result means that neither MTBC DNA nor the sample processing control was detected. An indeterminate result indicates a failure in the BD MAX system, while an incomplete result means that the assay run was not fully completed(10). All the isolates showing unreportable, MTB unresolved, or indeterminate results were retested using 2.5 mL of the mixture of 500 µL of the inactivated cultures + 2.5 mL of STR (1:5 dilution).

#### Gene sequencing

Following the methodology described by Pinhata et al.(13), isolates exhibiting discordant results between BD MAX and MTBDR*plus* were subjected to Sanger gene sequencing. Additionally, isolates showing inferred mutations by MTBDR*plus* were also sequenced.

The *inhA* promoter was amplified and sequenced with primers inhA-1 and inhA-2 (positions −168 to 80 in reference to start codon)(21). The *katG* gene was sequenced with forward and reverse primers katG-P4, katG-P5, katG-P6, katG-P7 and katG-P8 (positions −135 to 2202 of *katG* plus 431 nucleotides after the end of the gene)(22). Primers rpoB-1 and rpoB-2 were used to amplify and sequence a 350-bp fragment of *rpoB* encompassing the RRDR (positions 1184 to 1533 from the start codon)(23). All amplification reactions were carried out using GoTaq G2 Flexi DNA polymerase kit and dNTP mix (Promega, Madison, WI, USA). Each PCR reaction included 1× colorless buffer, 200 µM dNTP mix, 1.5 mM MgCl_2_ for *inhA* promoter and *katG*, 1.75 mM MgCl_2_ for *rpoB*, 5 pmol of primers for *inhA* promoter and *katG*, 10 pmol of primers for *rpoB*, 0.75 U of Taq DNA polymerase, about 50 ng (2.6 μL) of DNA template and nuclease-free water (Qiagen, Valencia, CA, USA) for a final volume of 25 μL. Amplification comprised a denaturation step at 94°C for 5 min, 40 cycles of 94°C for 1 min, 64°C for *inhA* promoter (62°C for *katG*, 60°C for *rpoB*) for 1 min, and 72°C for 1 min. The final extension occurred at 72°C for 10 min. The amplicons were purified with ExoSAP-it (Affymetrix, USA) and sequenced with ABI 3130×L Genetic Analyzer and the BigDye Terminator version 3.1 Kit (Applied Biosystems, USA). Sequences were analyzed using BioEdit v7.2.5 software and the MUBII-TB-DB and BLAST tools(24).

#### Phenotypic DST by BACTEC MGIT 960

Isolates with mutations not listed in the WHO catalogue, as well as those with uncertain or interim associations with resistance, were subjected to pDST to RIF and/or INH in critical concentrations of 0.5 µg/mL and 0.1 µg/mL, respectively, using the BD BACTEC™ MGIT™ 960 IR Kit (Becton, Dickinson and Company, USA)(25).

#### Data analysis

The BD MAX performance was evaluated regarding MTBC identification and the detection of resistance to RIF and INH, using as reference methods MTBDR*plus* and, when applicable, gene sequencing. The sensitivity, specificity, and accuracy of the BD MAX were calculated(26).

## Results

### Retrospective BD MAX testing with NTM and MTBC clinical isolates with known mutations

All the NTM and *N. asteroides* cultures were negative for the MTBC by the BD MAX assay. Results for the MTBC isolates with known mutations are shown in Table 2. All the discordant results were repeated.

As shown in Table 2, BD MAX results were all positive for the MTBC. Regarding *rpoB,* 23 of the 29 isolates tested presented mutations, three of them silent (T427T, F433F and L443L – all interpreted as RIF-R by the BD MAX). Nineteen of the 20 isolates with non-silent *rpoB* mutations were interpreted as RIF-R by the BD MAX, including isolates with double mutations. One isolate with *rpoB* S450W high-confidence mutation was initially interpreted as RIF susceptible by the BD MAX (after repetition of the assay, the mutation was detected). All the six isolates with wild-type *rpoB* results were interpreted as RIF susceptible by the BD MAX (Table 2).

Regarding *katG* and *inhA,* BD MAX detected INH resistance in 14 of the 15 isolates with mutations. The only isolate with mutation interpreted as INH susceptible by BD MAX showed a heteroresistance profile in *inhA* (T-8C+WT). The remaining 14 isolates wild-type in both *katG* and *inhA* were correctly interpreted as INH susceptible by BD MAX (Table 2).

### Prospective BD MAX testing on suggestive*-*MTBC clinical isolates

We prospectively analyzed 1,440 cultures presumptively identified as MTBC based on colony morphology, received at our laboratory between June and August 2023. Each isolate was tested using both the MTBDR*plus* assay and the BD MAX system, and the results were compared.

Among the 1,440 isolates tested, MTBDR*plus* identified 1,409 as MTBC. Of these, initial testing by BD MAX detected 1,402 (99.5%) as MTBC-positive, six (0.4%) as MTBC-negative, and one (0.07%) as indeterminate (due to a system error). Among the 31 isolates identified as non-MTBC by MTBDR*plus*, initial testing by BD MAX identified 30 (96.8%) isolates as non-MTBC. Upon repeating BD MAX testing for isolates with discordant and indeterminate results, three of the six MTBC isolates initially identified as negative and the indeterminate isolate were subsequently confirmed as MTBC-positive. Of the three isolates that turned MTBC-positive upon BD MAX retesting, two were initially MTBC low positive (one of which was TUB band positive but inconclusive for drug resistance by LPA, i.e., had low bacterial load). Additionally, the non-MTBC isolate incorrectly identified as MTBC by BD MAX (the first result was MTBC low positive) was detected as MTBC-negative on retesting. No isolates showed incomplete results by BD MAX.

Overall, the BD MAX system demonstrated a sensitivity of 99.8% (1,406/1,409), a specificity of 100% (31/31), and an accuracy of 99.8% (1,437/1,440) for MTBC detection in culture isolates.

Results of both tests, including repetitions when necessary, for the detection of MTBC drug resistance are presented in Table 3. The BD MAX was concordant for 1,307 (99.2%) among the 1,318 wild-type isolates detected by the MTBDR*plus*. Eleven wild-type isolates showed discordant results on BD MAX. Among these, one was reported as RIF-R, one as INH-R, and three as non-MTBC (although these patients were diagnosed and treated for TB). Four isolates were reported as MTBC low positive, including one with a low bacterial load on LPA. Additionally, two isolates had an unreportable INH result with RIF wild-type; both were initially classified as MTBC low positive before retesting.

**Table 3.** Results of GenoType MTBDR*plus* and BD MAX MDR-TB for *Mycobacterium tuberculosis* complex isolates tested prospectively.

| LPA GenoType MTBDR<sub>plus</sub> |  |  |  |  |  |  |
| --- | --- | --- | --- | --- | --- | --- |
| BD MAX MDR-TB | Wild-type | RIF-R | INH-R | MDR | MTBC positive,<br>resistance inconclusive | Total |
| Wild-type | 1307 | 2<sup>b</sup> | 1 | 1 | - | 1311 |
| RIF-R | 1 | 40 | - | - | - | 41 |
| INH-R | 1 | - | 30 | 1<sup>c</sup> | - | 32 |
| MDR | - | - | - | 13 | - | 13 |
| Non-MTBC | 3 | - | - | - | - | 3 |
| MTBC low positive | 4 | - | - | - | 2 | 6 |
| INH UNR | 2<sup>a</sup> | - | - | 1<sup>a</sup> | - | 3 |
| **Total** | **1318** | **42** | **31** | **16** | **2** | **1409** |

RIF-R: rifampicin-resistant; INH-R: isoniazid-resistant; MDR: multidrug-resistant; MTBC low positive: *M. tuberculosis* complex DNA detected, resistance metrics not measurable due to low bacterial load; INH UNR: BD MAX error INH resistance unreportable (MTBC DNA detected but INH resistance metrics not measurable; either the *katG* or *inhA* promoter probe failed to give a signal with the other signal presenting as wild type); MTBC positive, resistance inconclusive: TUB band positive but the genes' bands were faint due to low bacterial load; <sup>a</sup>RIF susceptible; <sup>b</sup>1 isolate with a heteroresistant profile in *rpoB* by LPA; <sup>c</sup>RIF unreportable: MTBC DNA detected but RIF resistance metrics not measurable (a single *rpoB* probe failed to give a signal with the remaining *rpoB* probes presenting as wild-type signals)

Concordance between the tests for RIF-R detection was 95.2% (40/42), and among the two discordant isolates, one had low bacterial load on LPA (*rpoB* WT6, WT7, WT8 bands absent), while the other had a heteroresistant profile on *rpoB* by LPA (WT+S450L) and was not retested by BD MAX (Table 3).

Of the 31 isolates identified as INH-R by LPA, 30 (96.8%) were concordant on BD MAX. The single discordant isolate lacked the *inhA* WT2 band on LPA but was reported as wild-type by BD MAX. Among the 16 MDR isolates detected by LPA, 13 (81.3%) showed concordance on BD MAX. Of the three discordant MDR isolates, one was identified as wild-type by BD MAX but exhibited a heteroresistant profile on LPA (*rpoB* WT+D435V and *katG* WT+S315T1) and was not subjected to BD MAX retesting; another was classified as INH-R with unreportable RIF by BD MAX, showing the absence of *rpoB* WT2, WT3 and WT4 bands and the *katG* S315T1 mutation on LPA; and the third isolate, which lacked the *rpoB* WT7 band and showed *katG* locus control and WT bands as negative, was INH unreportable and RIF wild-type on BD MAX. The two MTBC isolates with inconclusive drug resistance results on LPA showed a MTBC low positive result on BD MAX, indicating the cultures had low bacterial load (Table 3).

The isolates with discordant results, excluding the non-MTBC and MTBC low positive isolates, were further analyzed using Sanger sequencing, as detailed in Table 4. Among the isolates identified as wild-type by LPA, the RIF-R and the INH-R isolates detected by BD MAX were confirmed to be heteroresistant through sequencing, demonstrating that BD MAX had greater sensitivity for mutation detection compared to LPA in these cases. Of the two isolates with INH unreportable results and RIF wild-type by BD MAX but identified as wild-type by LPA, sequencing revealed that one was *katG* heteroresistant (mutation missed by LPA), while the other was wild-type for both *katG* and *inhA*, in concordance with the LPA.

**Table 4.**
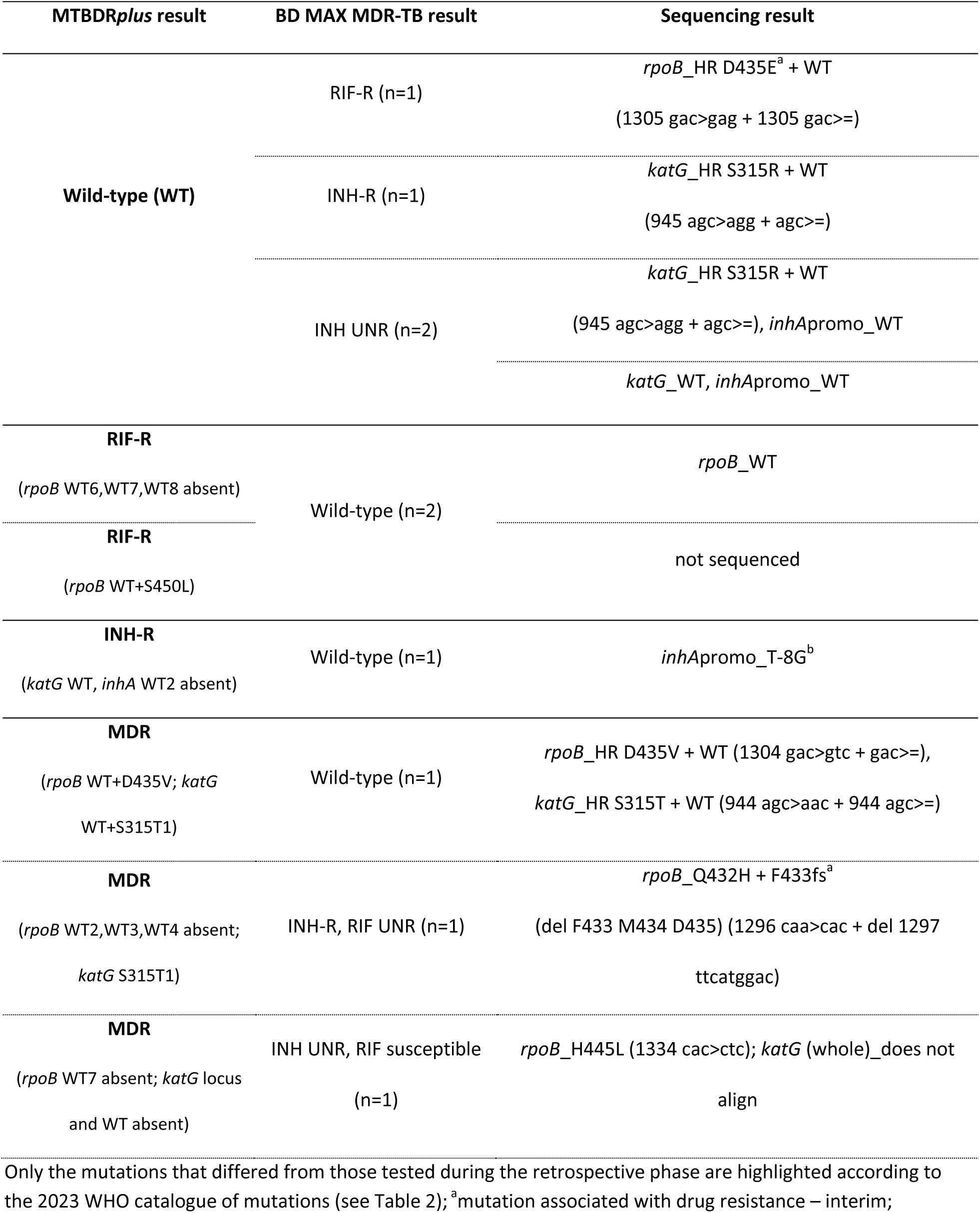
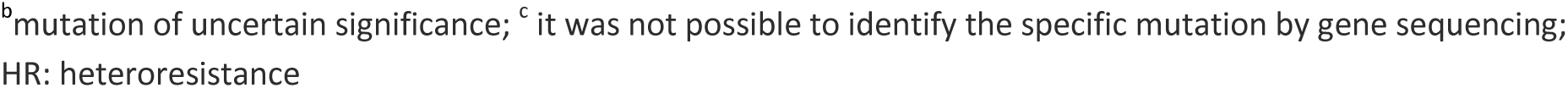
Gene sequencing results of *Mycobacterium tuberculosis* complex isolates showing resistance discordances between GenoType MTBDR*plus* and BD MAX MDR-TB assays.

For the two isolates classified as RIF-R by LPA but wild-type by BD MAX, sequencing confirmed that one was indeed wild-type for the *rpoB* gene. The other isolate was not sequenced due to an *rpoB* heteroresistant profile detected by LPA. The wild-type isolate correctly detected by BD MAX showed no WT6,7,8 bands on LPA, indicating, by our experience, a low bacterial load in the culture, leading to false-resistant results. This was corroborated by the patient’s RIF-susceptible result obtained through Xpert Ultra. Regarding the single INH-R discordant isolate, BD MAX failed to identify the *inhA* promoter T-8G mutation, which was identified by sequencing, in the isolate with absence of the WT2 band on LPA (Table 4).

Among the MDR isolates that showed discordances by BD MAX, one was classified as wild-type by BD MAX but exhibited a heteroresistant profile on LPA for both *rpoB* and *katG*, which was confirmed by sequencing; another isolate was INH-R and RIF unreportable by BD MAX, with sequencing revealing a single nucleotide polymorphism (SNP) followed by a deletion in *rpoB*, which is an infrequent mutation classified as associated with RIF resistance-interim. For the third discordant MDR isolate, which was detected as RIF-susceptible and INH unreportable by BD MAX, sequencing identified the presence of the *rpoB* H445L borderline mutation, associated with RIF resistance, while its *katG* sequence did not align with the reference, indicating a deletion in *katG* gene, as there was no presence of *katG* locus and WT bands on LPA (Table 4).

## Discussion

The BD MAX system represents a new generation of closed, automated assays using medium-to high-throughput platforms, enabling rapid detection of MTBC and resistance to RIF and/or INH. This is the first study evaluating the off-label use of the BD MAX MDR-TB platform in clinical isolates under routine conditions in a high TB burden setting. The results demonstrate the BD MAX system’s robust sensitivity, specificity, and accuracy in both retrospective and prospective analyses.

Most studies evaluating the BD MAX system were conducted on clinical samples(11,12,27–31), as this test is primarily recommended for a rapid initial diagnosis of TB and drug resistance detection. A multicenter study evaluating sputum samples found that the BD MAX assay on raw sputum had a sensitivity of 93% and a specificity of 99% for TB detection compared to culture and/or Xpert. For RIF resistance, BD MAX demonstrated a sensitivity of 90% and a specificity of 98% when compared to pDST and conventional genetic sequencing. In detecting INH resistance, the BD MAX assay showed a sensitivity of 82% and a specificity of 100% compared to pDST(12).

As a reference laboratory that exclusively processes mycobacterial cultures and does not perform initial TB diagnosis, we have successfully validated the BD MAX system for use with clinical isolates regarding the LoD of the assay, incubation protocols with the STR reagent, and the optimal ratio of inactivated culture samples to STR. According to our results, the LoD for MTBC detection in an H37Rv inactivated culture was 100 CFU/mL, while the LoD for drug resistance-associated mutations was lower, 1,000 CFU/mL. Beutler et al. also observed a lower LoD for drug resistance mutations detection when compared to MTBC detection for BD MAX (32).

In the retrospective testing, BD MAX accurately excluded all NTM and *Nocardia asteroides* cultures, confirming its high specificity for MTBC. A previous study focusing on BD MAX sensitivity with paucibacillary, smear-negative samples and samples growing NTM obtained 100% specificity for pulmonary samples growing NTM on culture(27). In Brazil, NTM have been increasingly isolated from clinical specimens and identified as causative agents of opportunistic infections, thus diagnostic assays must reliably distinguish between infections caused by MTBC and NTM. BD MAX demonstrated strong performance in this regard, effectively differentiating between these pathogens.

For MTBC isolates with known mutations tested retrospectively, BD MAX detected all isolates as MTBC, and most isolates showed concordance in terms of mutations detection in both RIF and INH. This is in accordance with previous studies, which observed BD MAX’s high sensitivity and specificity for detection of mutations in the target genes for MTBC isolates, pulmonary and extrapulmonary samples(11,27,30,33).

The MTBDR*plus* assay has probes that identify the most common high-confidence mutations encountered in *rpoB, katG* and *inhA* promoter. In contrast, the BD MAX system does not provide information about the specific mutation detected. Instead, it indicates whether a mutation is present and identifies the associated gene where the mutation occurs. This design ensures rapid results but lacks the granularity to pinpoint precise nucleotide or codon changes, which may be necessary for detailed resistance profiling or epidemiological studies.

In this study, silent mutations in *rpoB* were interpreted as RIF-R by the BD MAX, highlighting its high sensitivity to any *rpoB* alterations, which may lead to false RIF-R results in some cases. These results might, therefore, be confirmed by use of other genotypic tests, such as sequencing or pDST to avoid unnecessary treatment in case of silent mutations and to detect borderline mutations often causing low-level resistance(27). Notably, in our retrospective analysis, BD MAX demonstrated a strong capacity to detect isolates with double mutations, as well as mutations in isolates showing heteroresistance, except for one isolate with the T-8C+WT profile in *inhA* promoter.

MTBDR*plus* is routinely performed in our laboratory, but it is a labor-intensive technique that requires multiple dedicated rooms. DNA extraction is conducted manually in batches of 47 isolates plus one negative control, and the PCR plate is also prepared manually. Hybridization of the 48 samples is carried out using the GT-Blot 48 equipment, with results analyzed using the GenoScan scanner and software. The entire process takes three days: one day for DNA extraction and PCR, another for hybridization, and a third for reading the results after the strips have adequately dried. In contrast, the BD MAX platform is significantly simpler to operate. According to our validated protocol, isolates are first inactivated by thermal lysis, then mixed with the STR reagent, shaken vigorously, and incubated for five minutes at room temperature. The prepared mixture is then ready to be transferred directly to the BD MAX equipment, and all steps, including result interpretation, can be completed on the same day. In our workflow, team members work 4-hour daily shifts, so it would be possible to test and release results for 24 samples per day by BD MAX, as each run takes less than 4 hours. While the BD MAX has a lower simultaneous testing capacity of 24 samples, it offers a significant advantage by providing same-day results, unlike the MTBDR*plus*, which requires three days to complete the process for 47 samples.

In addition to testing NTM, frequent and less frequent mutations found routinely in MTBC clinical isolates in our setting, we also tested silent *rpoB* mutations to evaluate BD MAX performance. Another key strength of our study is the large number of isolates tested prospectively under real-world routine conditions. This included isolates cultured on solid media or in MGIT with varied ages, as well as those with low bacterial loads or contamination by other microorganisms. Testing a diverse and adequate sample size with various mutations is essential for obtaining precise diagnostic accuracy estimates.

The prospective analysis of 1,440 clinical mycobacterial isolates further validated the BD MAX system’s effectiveness in our routine. The system achieved a sensitivity of 99.8% and specificity of 100% for MTBC detection, with discrepancies primarily related to low bacterial loads or heteroresistant profiles. This level of performance aligns well with the MTBDR*plus* assay, the test used in our laboratory, but with the added advantage of automation and reduced hands-on time. In our workflow, inconclusive results obtained by LPA are repeated. Similarly, if BD MAX were used instead of LPA, MTBC low positive, indeterminate, and unreportable results would also undergo repetition before releasing the final report. BD MAX’s ability to correctly reclassify discordant and indeterminate results upon retesting underscores its reliability in routine diagnostic settings.

In this study, we observed a lower rate of indeterminate or unreportable results compared to other studies conducted directly on clinical samples, as the one by Zimmermann et al. (2018) who reported 8.9% indeterminate RIF/INH results using BD MAX on 518 clinical samples(34). This difference is likely because we performed BD MAX testing on clinical isolates, which contain significantly higher concentrations of microorganisms than clinical specimens.

Sequencing of discordant isolates tested prospectively by LPA and BD MAX revealed that BD MAX detected heteroresistance in two cases considered wild-type by LPA, one RIF-R (presenting the heteroresistant *rpoB* D435E+WT profile) and the other INH-R (presenting the heteroresistant *katG* S315R+WT profile). However, both BD MAX and LPA failed to detect the same *katG* S315R mutation in another heteroresistant isolate, which was INH unreportable by BD MAX and wild-type by LPA. The other isolate classified as INH unreportable by BD MAX and wild-type by LPA was confirmed as wild-type by sequencing. Notably, both isolates reported as INH unreportable yielded MTBC low positive results on the initial BD MAX run, suggesting an association between unreportable resistance results and low bacterial load, as reported by Hofmann-Thiel et al. (2020)(27).

Based on our findings, the BD MAX assay successfully detected a wild-type isolate wrongly classified as RIF-R by LPA, attributed to the absence of WT6, WT7, and WT8 bands—an outcome typically associated with a low bacterial load in the sample. This highlights the lower LoD of BD MAX compared to LPA. According to the manufacturers, the BD MAX assay has a LoD of 0.25 CFU/mL for detecting MTBC in sputum specimens(10). In contrast, the LoD for LPA is significantly higher, at 160 CFU/mL for sputum samples(18).

The BD MAX assay missed certain specific mutations detected by LPA, including an *inhA* promoter mutation (T-8G) and a heteroresistant MDR isolate with an *rpoB* D435V+WT and *katG* S315T+WT profile. Interestingly, this patient had an Xpert Ultra RIF-susceptible result. Additionally, BD MAX failed to resolve some resistance patterns detected as inferred mutations by LPA. For example, an isolate with double mutations in *rpoB* (Q432H+F433fs) was classified as RIF unreportable, while another isolate with *rpoB* H445L borderline mutation and a deletion in *katG* was detected as RIF wild-type and INH unreportable. These findings highlight limitations in BD MAX’s mutation coverage and its ability to interpret complex resistance profiles.

Our study has some limitations. Mycobacterial cultures tested prospectively were received from many TB laboratories and varied in age and in bacterial load, potentially affecting the homogeneity of testing conditions. Although culture-to-STR volume ratios were standardized, variability in culture conditions might still influence results. Furthermore, gene sequencing by Sanger could not specifically identify the mutation present in the MDR isolate with no *katG* locus and WT bands in the LPA. In this case, whole genome sequencing would elucidate the specific mutation.

In conclusion, the BD MAX platform proves to be a valuable tool for TB diagnostics, particularly in high-burden settings, offering reliable MTBC detection and resistance profiling in culture samples. Its performance supports its integration into routine diagnostics of high-volume reference laboratories, with sequencing or pDST recommended as a confirmatory method for complex or inconclusive cases.

## Data Availability Statement

The DNA sequences of all the sequenced isolates were deposited in GenBank (http://www.ncbi.nlm.nih.gov/) under the following identification: PRJNA888434.

## Acknowledgments

We would like to thank André Luis Santos at Becton, Dickinson and Company for the provision of funding and supplies for this study. Becton, Dickinson and Company did not have a role in study design, procedures, analysis or writing of the report.

The authors thank the technical assistance and support provided by the staff of Núcleo de Tuberculose e Micobacterioses, namely Andréia Rodrigues de Souza, Aparecida Andrade Pereira, Flávia de Freitas Mendes, Juliana Failde Gallo, Sonia Maria da Costa and Vera Lucia Maria da Silva, and Centro de Bacteriologia of Adolfo Lutz Institute.

## Author contributions

Angela Pires Brandao: Data curation, Formal analysis, Investigation, Writing – review and editing, Visualization, Supervision. Fabiane Maria de Almeida Ferreira: Investigation. Fernanda Cristina dos Santos Simeao: Investigation. Lucilaine Ferrazoli: Writing – review and editing. Erica Chimara: Funding acquisition, Writing – review and editing. Rosangela Siqueira de Oliveira: Investigation, Supervision, Writing – review and editing. Juliana Maira Watanabe Pinhata: Conceptualization, Data curation, Formal analysis, Funding acquisition, Investigation, Methodology, Project administration, Supervision, Writing – original draft, Writing – review and editing.

